# Coordinated regulation of gene expression and microRNA changes in adipose tissue and circulating extracellular vesicles in response to pioglitazone treatment in humans with type 2 diabetes

**DOI:** 10.1101/2021.10.30.21265710

**Authors:** Yury O. Nunez Lopez, Anna Casu, Zuzana Kovacova, Alejandra Petrilli, Olga Sideleva, William G. Tharp, Richard E. Pratley

**Author notes:** **Address correspondence to:** Richard E. Pratley, MD. AdventHealth Translational Research Institute, 301 East Princeton Street, Orlando, FL, 32804.

## Abstract

Pioglitazone, a PPARγ agonist, is used to treat type 2 diabetes (T2D). PPARγ is highly expressed in adipose tissue (AT), however the effects of pioglitazone to improve insulin sensitivity are also evident in other tissues. We hypothesized that pioglitazone modifies the cargo of circulating AT-derived extracellular vesicles (EVs) to alter interorgan crosstalk. We tested this in a 3-month trial in which 24 subjects with T2D who were well-controlled with diet/exercise or metformin were randomized to treatment with either pioglitazone 45 mg/day or placebo (NCT00656864). Levels of 42 adipocyte-derived EV-miRNAs were measured in plasma EVs. Levels of 5 miRNAs (i.e., miR-7-5p, miR-20a-5p, miR-92a-3p, miR-195-5p, and miR-374b-5p) were significantly downregulated in EVs in response to pioglitazone treatment relative to placebo. However, the opposite occurred for miR-195-5p in subcutaneous AT from the same participants. Changes in miRNA expression in EVs and AT correlated with changes in suppression of lipolysis and improved insulin sensitivity, among others. DICER was downregulated and exosomal miRNA sorting-related genes YBX1 and hnRNPA2B1 displayed a trend toward downregulation in AT. Furthermore, analysis of EV-miRNA targeted genes identified a network of overtargeted transcripts that changed in a coordinated manner in AT. Collectively, our results suggest that some beneficial pharmacologic effects of PIO are mediated by adipose-specific miRNA regulation and exosomal/EV trafficking.

**Disclosure summary:** This study was funded by program funds granted to REP by the AdventHealth Translational Research Institute. The clinical trial was supported by an investigator initiated grant to REP from Takeda Pharmaceuticals North America. REP reports grants from Hanmi Pharmaceutical Co.; grants from Janssen; consulting fees from Merck; grants, speaker fees and consulting fees from Novo Nordisk; consulting fees from Pfizer; grants from Poxel SA; grants and consulting fees from Sanofi; consulting fees from Scohia Pharma Inc.; consulting fees from Sun Pharmaceutical Industries. AC reports consulting fees from GlaxoSmithKline. Honoraria and fees for REP’s and AC’s services were paid directly to AdventHealth, a nonprofit organization. No other potential conflicts of interest relevant to this article were reported.

## Introduction

Exosomes and microvesicles (herein referred to as extracellular vesicles/EVs) are small particles, ranging in size from 30-400 nm, that are released from most cell types(1). EVs contain a variety of cargo molecules, including microRNAs, mRNAs, proteins and lipids, reflecting the cell of origin as well as its physiological state. Adipose tissue (AT) has been identified as a major source of circulating EV microRNAs (EV-miRNAs)(2). EVs and EV-miRNAs mediate crosstalk between cells within and between tissues as well as between organs in both health and disease, including diabetes(3). EVs are, thus, potential effectors of metabolic responses to pharmacologic agents for the treatment of diabetes.

Pioglitazone (PIO), an approved treatment for type 2 diabetes (T2D), is a potent agonist of peroxisome proliferator-activated receptor-γ (PPARγ), which is predominantly expressed in adipose tissue (AT) and is a master regulator of adipogenesis(4). PPARγ agonism has been shown to alter cancer derived EV-miRNA (5), but it is not known whether pioglitazone treatment in people with T2D exerts any pharmacologic effect via regulation of AT EV miRNA. To a lesser extent, PIO also activates PPARα, which is expressed in many metabolically active tissues, is required for ketogenesis in the liver, and reduces inflammation by repressing endothelial TNFα-induced genes(6). Treatment with PIO improves insulin sensitivity and hyperglycemia in patients with T2D and prevents major adverse cardiovascular events in patients with insulin resistance and T2D (7, 8). Although PIO treatment induces weight gain, a shift from visceral to subcutaneous fat distribution occurs and this shift associates with improvements in insulin sensitivity in the liver and peripheral tissues(9). Interestingly, pioglitazone treatment was reported to normalize miR-29 levels in the livers of Diet-Induced Obesity (DIO) mouse model and in the Zucker Diabetic Fatty (fa/fa) rats (10). miR-29 is also known for negatively regulating insulin signaling in adipocytes, but a definitive mechanism has not been elucidated (11, 12). We hypothesized that pioglitazone exerts effects via the modulation of AT-derived EV cargo (i.e., EV-miRNAs) and that these changes would be related to changes in metabolic function in subjects with T2D.

## Results and Discussion

To test our hypothesis, we used archived plasma and subcutaneous adipose tissue samples from a 3-month randomized, controlled trial (NCT00656864)(13). Participants included 24 subjects with T2D who were well-controlled (HbA1c ≤ 7.0%) with diet/exercise or metformin. Participants were randomized to treatment with either pioglitazone 45 mg/day (PIO, n=12) or placebo (PLA, n=12). The two groups were balanced with respect to most baseline clinical measures (Table 1). As reported in the parent study, treatment with pioglitazone increased weight and BMI and was associated with significant improvements in insulin sensitivity and HbA1c levels(13).

**Table 1.** Clinical characteristics of the study cohort.

|  | Level | Pre-treatment (Baseline) |  | *P* |
| --- | --- | --- | --- | --- |
|  |  | Placebo (n=12) | Pioglitazone (n=12) |  |
| Gender (%) | F | 5 (41.7) | 7 (58.3) | 0.683 |
|  | M | 7 (58.3) | 5 (41.7) |  |
| Metformin (%) | N | 4 (33.3) | 5 (41.7) | 1 |
|  | Y | 8 (66.7) | 7 (58.3) |  |
| Age |  | 61.50 [56.00, 66.00] | 62.50 [48.50, 65.25] | 0.885 |
| BMI (kg/m<sup>2</sup>) |  | 31.65 [26.12, 34.92] | 34.65 [28.35, 39.43] | 0.623 |
| Height (cm) |  | 173.75 [158.50, 181.88] | 165.50 [164.12, 173.12] | 0.773 |
| Weight (kg) |  | 87.80 [82.92, 96.00] | 91.75 [88.17, 100.05] | 0.236 |
| DEXA Percent Body Fat (%) |  | 37.30 [29.70, 46.05] | 34.20 [31.30, 45.85] | 0.854 |
| Waist circumference (cm) |  | 105.25 [99.38, 119.00] | 108.85 [102.75, 113.38] | 0.707 |
| Hematocrit (%) |  | 39.40 [36.72, 42.78] | 39.15 [36.38, 41.85] | 0.729 |
| Total Cholesterol (mg/dL) |  | 176.50 [168.50, 181.50] | 176.00 [145.25, 189.00] | 0.751 |
| Triglyceride (mg/dL) |  | 132.50 [83.75, 193.25] | 125.50 [99.00, 203.00] | 0.773 |
| HDL (mg/dL) |  | 41.00 [38.75, 43.00] | 46.50 [42.75, 51.25] | 0.077 |
| LDL (mg/dL) |  | 97.00 [89.00, 108.50] | 94.50 [73.75, 100.25] | 0.479 |
| FFA (mmol/L) |  | 0.55 [0.34, 0.61] | 0.48 [0.45, 0.64] | 0.626 |
| K<sub>FFA</sub> (% x min<sup>-1</sup>) |  | 5.73 [3.37, 7.19] | 4.99 [4.05, 6.75] | 0.922 |
| Serum TNFα (pg/ml) |  | 4.23 [3.37, 5.51] | 4.15 [4.02, 4.50] | 0.622 |
| Hemoglobin A1c (%) |  | 6.65 [5.97, 6.82] | 6.30 [6.10, 6.53] | 0.839 |
| Fasting Glucose (mg/dL) |  | 126.25 [94.67, 139.62] | 97.50 [92.04, 105.38] | 0.583 |
| Fasting Insulin (μIU/mL) |  | 12.10 [6.22, 19.35] | 4.26 [3.33, 10.59] | 0.488 |
| Fasting C peptide (ng/mL) |  | 2.73 [2.13, 3.44] | 1.88 [1.50, 3.03] | 0.947 |
| Si (min<sup>-1</sup> x (μIU/ml)<sup>-1</sup>) |  | 0.90 [0.66, 1.47] | 1.63 [1.20, 1.86] | 0.514 |
| Sg (min<sup>-1</sup>) |  | 0.01 [0.01, 0.02] | 0.02 [0.01, 0.02] | 0.514 |
| AIRg (μIU/ml x10min) |  | 76.01 [7.26, 512.89] | 68.34 [41.31, 132.31] | 0.935 |
| DI |  | 64.71 [28.24, 326.57] | 67.15 [40.23, 111.86] | 0.87 |

*Data presented as median with interquartile range in square brackets, otherwise specified.*

EVs were isolated from fasting plasma samples collected before and after the 12-week treatment period. The characterization of the EV preparations by transmission electron microscopy (TEM) and Western blot (Figure 1A,B) showed an enrichment in nanoparticles with sizes, shapes, and membrane protein markers that were compatible with those of exosomes and small microvesicles(1). In addition, these preparations were free of the non-exosomal marker calnexin (Figure 1B). Neither PIO nor PLA treatment significantly changed the average number of circulating EVs in the study subjects, as quantified by nanoparticle tracking analysis (NTA) and CD9 ELISA (Supplementary Figure SF1).

**Figure 1.**
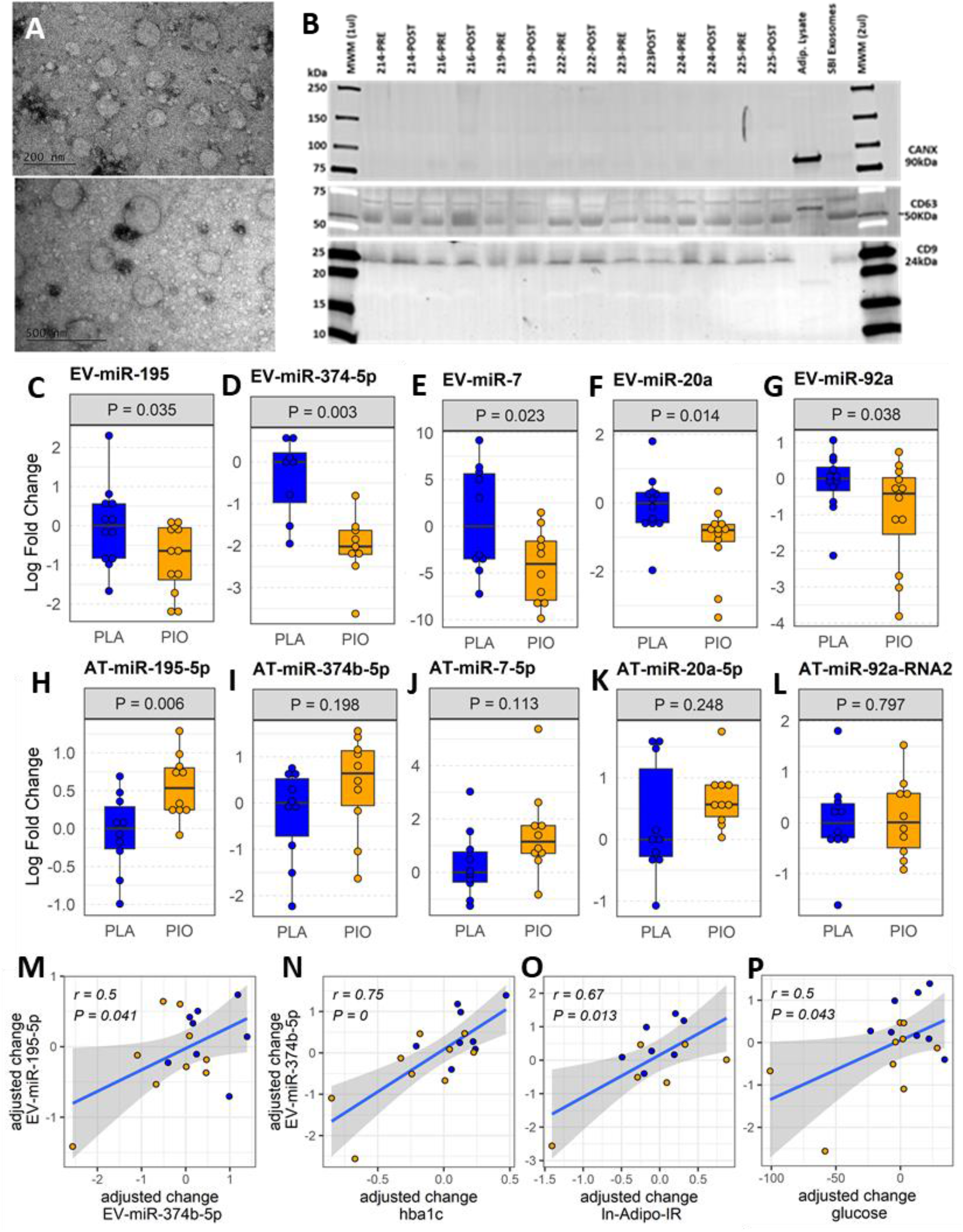
Profiling plasma EV-miRNAs and AT-miRNAs in subjects with T2D treated with pioglitazone (PIO, yellow color filling) or placebo (PLA, blue color filling). **(A)** Transmission electron microscopy. **(B)** Western blot for exosomal (CD9, CD63) and non-exosomal (CANX) protein markers. **(C-G)** Baseline-adjusted boxplots of five significantly downregulated miRNAs in circulating EVs from T2D subjects treated with PIO, compared to PLA. **(H-L)** Baseline-adjusted boxplots of miRNA expression in adipose tissue (AT) from T2D subjects treated with PIO, compared to PLA group. **(M-P)** Significant partial correlations between EV-miRNAs and relevant clinical measures.

### Five miRNAs were downregulated in circulating EVs after treatment with pioglitazone

The abundance of 42 miRNAs (Supplementary Table ST1) reported to be expressed in adipocyte-derived EVs(2) was profiled in EVs isolated from fasting plasma obtained before and after treatment. The levels of five circulating EV-miRNAs (*i.e*., EV-miR-374b-5p, EV-miR-195-5p, EV-miR-20a-5p, and EV-miR-7-5p, and EV-miR-92a-3p) were significantly downregulated (fold change<-1.5, *P*<0.05, FDR≤0.15) in response to PIO as compared to PLA treatment (Figure 1C-G, Supplementary Table ST2-A). Our statistical analysis adjusted for body weight and hematocrit measurements to account for the potential confounding effects of weight gain and increased blood volume known to be induced by pioglitazone. Interestingly, the change in EV-miR-374b-5p significantly correlated with the change in EV-miR-195b-5p (r=0.5, P=0.041, FDR=0.11, Figure 1M) and suggested coregulation of these EV-miRNAs. Importantly, the change in EV-miR-374b-5p strongly correlated with the change in HbA1c, fasting plasma glucose, and insulin resistance in the adipose tissue (r≥0.5, P<0.05, FDR<0.1, Figure 1N-P), among others. These correlations suggest that the changes in plasma EV-miRNAs might be functionally associated with the glycemic control exerted by the pioglitazone treatment.

### miR-195-5p is upregulated in subcutaneous adipose tissue after treatment with pioglitazone

Considering that AT is one of the main sites of pioglitazone action and that AT contributes a majority of EV-miRNAs into the circulation(2), we quantified the respective miRNA expression levels of the differentially expressed EV-miRNAs in subcutaneous AT samples from the same participants. In contrast to the downregulation of circulating EV-miRNAs, the expression of the corresponding miRNAs in AT (AT-miRNAs) was unchanged or increased in response to pioglitazone relative to placebo (Figure 1H-L, Supplementary Table ST2-B). Only miR-195-5p showed a statistically significant (P=0.006, FDR=0.03, Figure 1H-L) increase in AT and this change was significantly correlated with the change in waist circumference (r=0.76, P<0.018, FDR<0.18) and negatively correlated with the change in serum concentration of TNFα and in the insulin sensitivity index (Si) (-r>0.5, P<0.02, FDR<0.1) (Figure 2A-C). Additionally, the changes in AT-miR-195-5p, AT-miR-7-5p, and AT-miR-20a-5p expression levels were strongly correlated (all r>0.65, P<0.01, FDR<0.025, Figure 2D-F), suggesting coregulated transcriptional mechanisms for these AT-miRNAs. Similarly, AT-miR-7-5p and AT-miR-20a-5p correlated with relevant clinical measures including levels of circulating LDL and Si, an index of insulin sensitivity (all r>0.55, P<0.02, FDR<0.05, Figure 2H-I). This suggests that the PIO-associated reduction in miR-195-5p in circulating EVs and corresponding increased expression in AT may contribute to improved inflammatory milieu, glycemic control, and insulin sensitivity in subjects with T2D. Interestingly, we previously reported that circulating miR-195-5p and miR-7-5p were reduced by short-term intensive insulin therapy in subjects with early T2D(14) and vitamin D supplementation in subjects with prediabetes(15), respectively. Collectively, these findings suggest general roles for EV-miR-195-5p and EV-miR-7-5p, in response to distinct anti-hyperglycemic treatments. Other authors have reported that miR-7-5p induces repression of β cell function and proliferation(16) and also plays a crucial role as PPARα-regulated mediator of lipid metabolism in hepatocytes, where it activates SREBP signaling and promotes lipid accumulation(17). Further supporting our findings, a study by Papi and colleagues reported that pioglitazone can abolish the capability of hypoxic breast cancer cell-derived exosomes to induce miRNA-mediated pro-inflammatory and pro-invasive pathways in surrounding tumor associated fibroblasts(5). We reason that, by analogy (i.e., modifying the miRNA content of secreted EVs), pioglitazone might mitigate the capacity of EVs to induce miRNA-mediated diabetogenic changes in metabolism.

**Figure 2.**
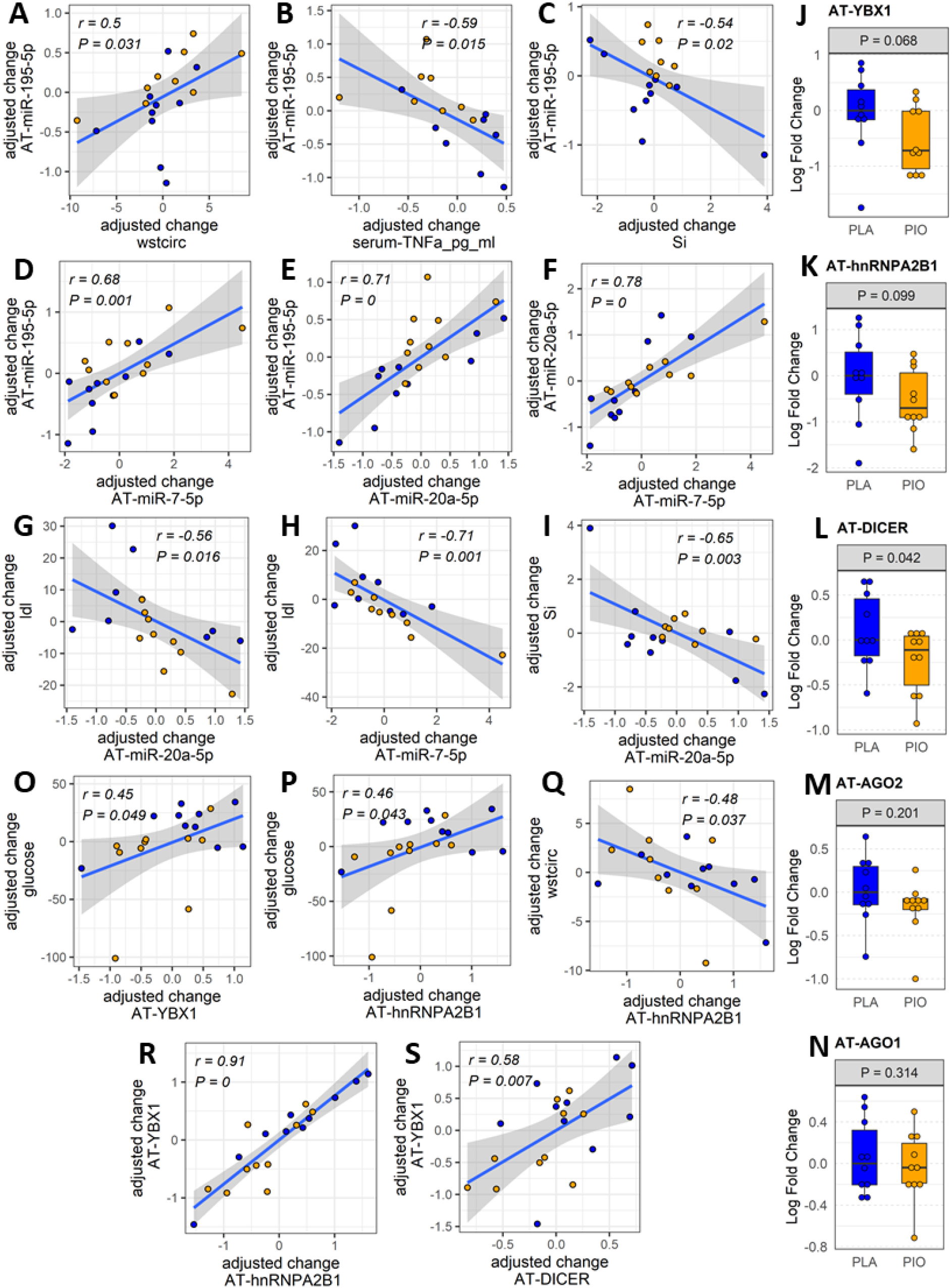
miRNAs and exosome sorting genes in AT. **(A-I)** Significantly correlated changes between DE AT-miRNAs and between DE AT miRNAs and relevant clinical and biochemical measures. **(J-N)** Expression of AT transcripts involved in miRNA exosomal sorting and miRNA biogenesis. **(O-S)** Significantly correlated changes between AT genes involved in miRNA exosomal sorting and relevant clinical measures. Yellow color filling for pioglitazone group, blue color filling for placebo group.

### Dicer, YBX1, and hnRNPA2B1 are downregulated in subcutaneous fat tissue from subjects with type 2 diabetes treated with pioglitazone

We reasoned that specific miRNAs upregulated in the AT (i.e., miR-195-5p) may be retained in the tissue by being diverted from the exosomal sorting pathway. To test this hypothesis, we evaluated potential causes of the reduced levels of circulating AT-derived EV-miRNAs in response to PIO. Using subcutaneous AT samples from the same participants, we quantified the expression of genes involved in miRNA biogenesis and/or sorting to the exosomal pathway (*i.e*., Dicer, Ago2, YBX1, hnRNPA2B1 and nSMase2/SMPD3)(18). Because levels of AT-miRNAs increased in the PIO-treated group, we hypothesized that biogenesis-related Dicer and/or Ago2 would be upregulated in the AT of these participants. Because the miRNA levels were reduced in the plasma EVs of the PIO-treated subjects, we further hypothesized that exosomal sorting-related YBX1, hnRNPA2B1, and/or SMPD3 would be downregulated in the AT. We observed a trend for downregulation of AT-YBX1 (P=0.068, FDR=0.19) and AT-hnRNP2AB1 (P= 0.099, FDR=0.24) in response to pioglitazone treatment (Figure 2J,K). However, contrary to our expectations, AT-Dicer was significantly downregulated in the PIO group (P=0.042, FDR=0.19, Figure 2L). No significant changes were observed in Ago2 expression or the related family member Ago1 (Figure 2M,N). These observations suggest that the upregulation of PIO-responsive miRNAs in AT does not occur because of increased Dicer/Ago2-mediated miRNA biogenesis but, at least in part, because of suppression of miRNA secretion via the exosome pathway. Supporting this, changes in AT-YBX1 and AT-hnRNPA2B1 significantly correlated with the pioglitazone-induced changes in fasting glucose and waist circumference (absolute |r|≥0.45, P<0.05, FDR<0.08, Figure 2O-Q, Supplementary Figure SF1). Positive correlations were also observed between the change in AT-YBX1 and AT-DICER as well as the change in AT-YBX1 and AT-hnRNPA2B1 (r≥0.58, P≤0.007, FDR≤0.03, Figure 2R,S). Collectively, these results suggest that PIO treatment alters the sorting of specific miRNAs into the EV compartment of AT in people with T2D, and this is related to the drug-induced improvements in glycemic control and body fat redistribution.

### A network of miRNA-overtargeted genes are downregulated in subcutaneous adipose tissue from subjects with type 2 diabetes treated with pioglitazone

To further evaluate the functional relevance of the altered EV-miRNAs in response to pioglitazone treatment, we built a network of validated miRNA-gene interactions that identified 96 transcripts as overtargeted (i.e. having a higher probability of miRNA-gene interaction) by the five differentially abundant EV-miRNAs (Figure 3A, *P*_*simulation*_=0.0052). The analysis shown in Figure 3B,C highlights the top cell compartments and KEGG pathways enriched in the overtargeted gene network, respectively. The caveola, a cell membrane compartment involved in cellular trafficking and signaling and in lipid turnover(19), was found to be a key EV-miRNA-targeted compartment in response to pioglitazone treatment. Deficiency in this compartment caused maladaptative autophagy that contributes to insulin resistance and altered adipocyte differentiation(20). On the other hand, consistent with a broad array of signaling involved with diabetes and its complications, the enrichment in overtargeted KEGG pathways highlighted miRNA-regulated pathways in cancer, resistance to endocrine therapy, FoxO signaling, PI3K/Akt signaling, and sphingolipid signaling, among others (Figure 3C). The recurrent link with cancer and the response to anticancer therapies suggests that pioglitazone affects conserved biochemical pathways contributing to both, diabetes and cancer. Specifically, the well-established role for phosphatidylinositol triphosphate (PI(3,4,5)P3) signaling to regulate insulin sensitivity and cell metabolism via modulation of Akt activity(21, 22), as well as tumor growth(23, 24) and migration of T lymphocytes via modulation of FoxO signaling(25, 26), among others, underscore the mechanistic impact of pioglitazone treatment in the modulation of metabolism, inflammation, and the tumor microenvironment. The miRNA-targeting of sphingolipid signaling is also important and likely related to the mechanisms by which pioglitazone restores the altered lipid metabolism in diabetes. Supporting this reasoning, profiling of circulating sphingolipids uncovered that pioglitazone treatment in people with metabolic syndrome induced a potent decrease in plasma ceramides and that some of those changes correlated with changes in adiponectin levels and insulin resistance(27, 28).

**Figure 3.**
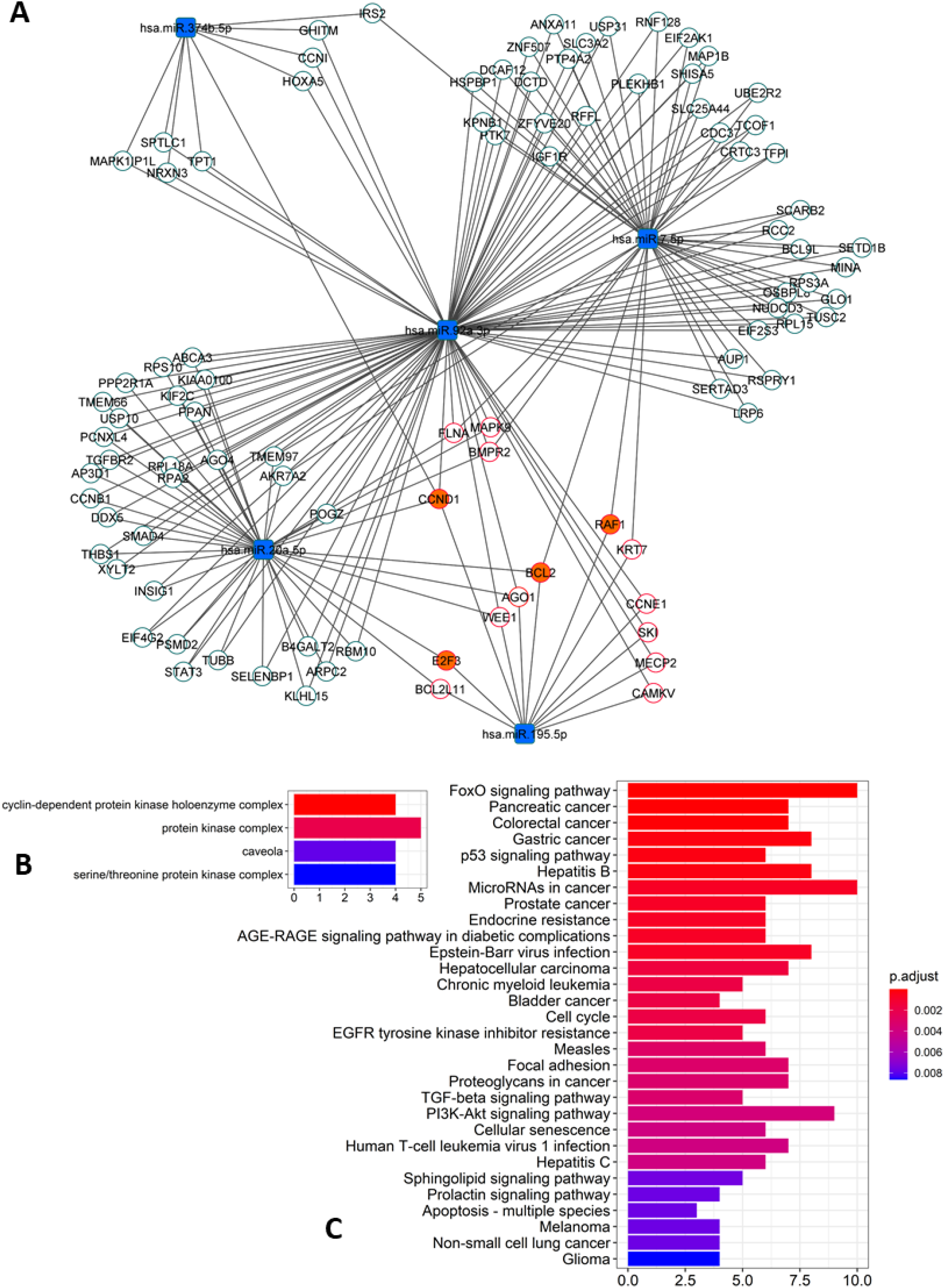
Network of overtargeted miRNA-mRNA interactions. **(A)** This particular type of network, only includes transcripts validated to interact with the differentially expressed (DE) miRNAs that were found to be targeted by more DE miRNAs than expected by chance (“ miRNA-overtargeted”). **(B)** Cellular compartments enriched among the annotations of the miRNA-overtargeted genes. **(C)** KEGG pathways enriched among the annotations of the miRNA-overtargeted genes.

To confirm the functional relevance of this miRNA-overtargeted gene network in a specific human tissue, we quantified the expression of overtargeted genes (specifically those validated as interacting with miR-195-5p as well as 3 other genes not targeted by miR-195-5p but central to the network by interacting with 3 of the 5 differentially abundant EV-miRNAs) in subcutaneous AT from the same study participants. Remarkably, the expression levels of 4 transcripts (*i.e*., RAF1, CCND1, BCL2, and E2F3) were significantly reduced in AT (Figure 4A-D) and strongly correlated with changes in targeting miRNAs (Figure 4G-J). These miRNA-overtargeted genes additionally displayed strong co-regulation in AT as indicated by highly significant correlated changes among each other (Figure 4K; Supplementary Table ST3). Their changes also significantly correlated with the change in differentially expressed Dicer and the exosomal miRNA-sorting genes YBX1 and hnRNPA2B1 in the AT (Figure 4K, Supplementary Table ST3). Most importantly, the change in these pioglitazone-associated AT genes significantly correlated with the change in key clinical measures of insulin secretion and function (fasting plasma insulin –FPI, insulin AUC, and insulin resistance in AT –Adipo IR), lipid metabolism (rate of free fatty acids disappearance –K_FFA, serum FFA concentration, HDL, and LDL), glycemic control (fasting plasma glucose –FPG), and body composition (waist circumference, tissue fat, free fat mass, and total mass) (absolute |r|≥0.45, P<0.05, FDR<0.1, Figure 4L-Z).

**Figure 4.**
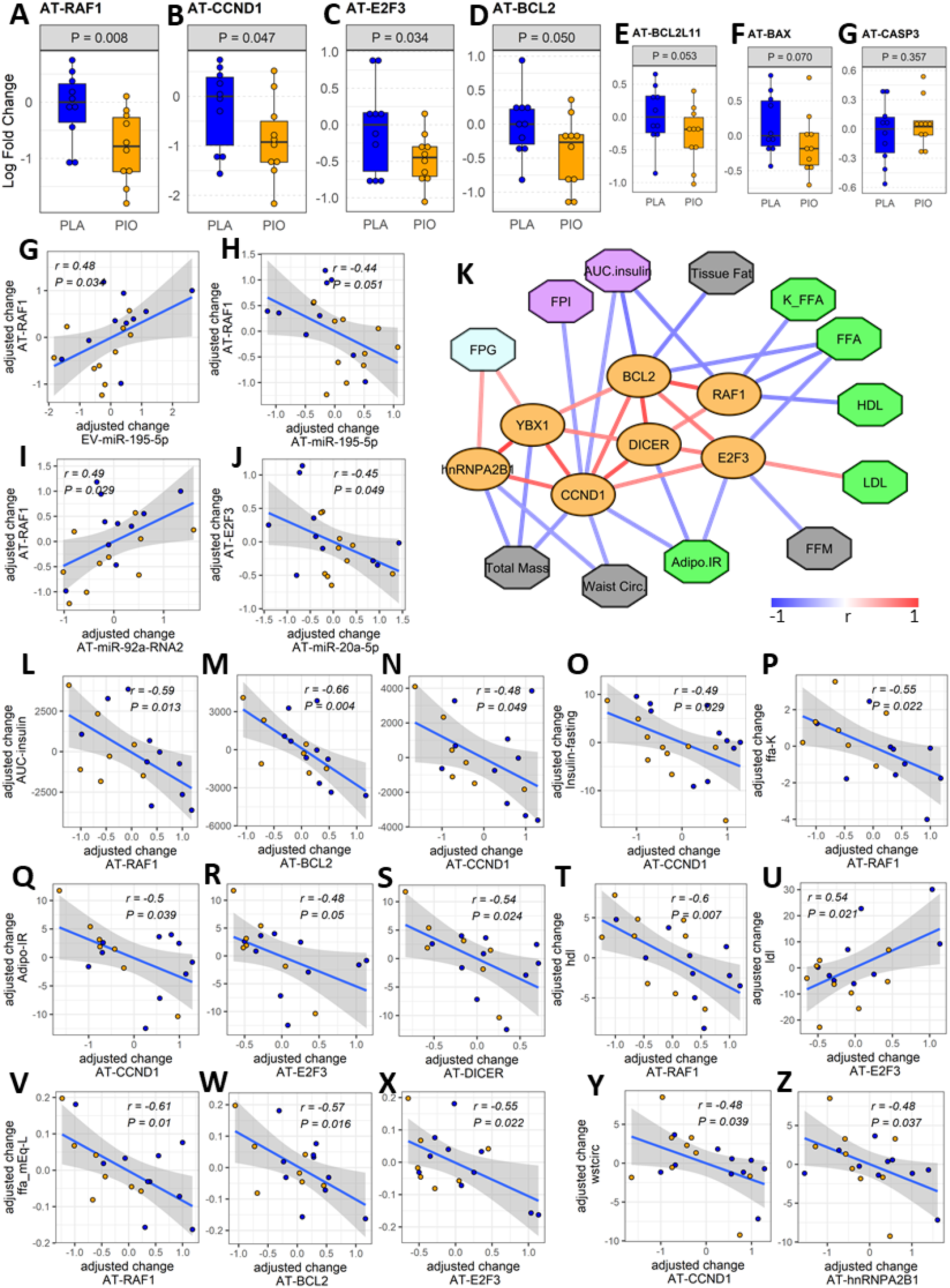
Expression and associations among AT genes found to be overtargeted by the DE EV miRNAs. **(A-E)** Profiles of differentially expressed overtargeted AT genes. **(F-G)** Profiles of pro-apoptotic BAX and CASP3 genes in AT. **(G-J)** Significant correlations between the change in DE overtargeted AT genes and the change in circulating DE EV miRNA and AT miRNAs. **(K)** Correlation network of significant (*P*<0.05, FDR<0.1) associations among AT genes involved in miRNA biogenesis/exosomal sorting, experimentally validated AT genes overtargeted by the DE EV-miRNAs (orange ovals), and clinical measures relevant to lipid metabolism (green octagons), body composition (grey octagons), glycemic control (cyan octagons), and β cell function (purple octagons). **(L-Z)** Correlation plots between the change in DE AT genes and the change in relevant clinical measures. Waist circ or wstcirc: waist circumference, FPG: fasting plasma glucose, FPI: fasting plasma insulin, AUC.insulin: insulin area under the curve, FFM: free fat mass, FFA: free fatty acids, K_FFA or ffa.K: rate of free fatty acid disappearance, LDL: low density lipoprotein, HDL: high density lipoprotein, Adipo.IR: insulin resistance in adipose tissue.

Of note, RAF1, the AT gene demonstrating the most significant downregulation in the subcutaneous AT of the PIO group (Figure 4A), is a known positive regulator of lipolysis in adipocytes(29). We reason that, by targeting RAF1, the pioglitazone-induced elevation of AT-miR-195-5p may enhance the suppression of lipolysis (also suggested earlier by the observed associations between the changes in AT-miRNAs and K_FFA and Adipo-IR), leading to weight gain, but also improved insulin sensitivity. Downregulation of RAF1 in AT could additionally contribute to antioxidant effects of pioglitazone with concomitant reduction of cellular stress(30). On the other hand, cyclin D1 (CCND1), another significantly downregulated miRNA-overtargeted gene in our study, was reported to inhibit mitochondrial function and size and its antisense inactivation shown to enhance oxidative glycolysis and lipogenesis(31). These findings suggest that CCND1 downregulation in AT may contribute to pioglitazone-induced weight gain. Interestingly, we now also report that changes in CCND1, RAF1, and E2F3 negatively correlate with the change in insulin AUC and in fasting plasma insulin (Figure 4K,N,O), which suggests an association between the expression of these pioglitazone-responsive miRNA-regulated genes in AT and insulin secretion from the β cells. This further suggests the existence of a modifiable adipose–pancreas interorgan communication axis that may involve circulating EVs with drug-inducible cargo modifications.

On the other hand, the significant downregulation of the miRNA-overtargeted anti-apoptotic gene BCL2 in AT from the PIO group seemed counterintuitive. However, in the context of a reduction of pro-apoptotic factors due to a presumable improvement of the inflammatory milieu, the compensatory reduction of anti-apoptotic factors seems plausible. Supporting this idea, the overtargeted pro-apoptotic gene BCL2L11 also demonstrated a trend toward downregulation (P=0.053, FDR=0.19, Figure 4E). To further assess our hypothesis, we additionally measured the expression of death-promoting BAX and CASP3 genes in AT. BAX demonstrated a trend (*P*=0.07, FDR=0.19) towards downregulation with no significant change in CASP3 levels (Figure 4F,G). Notably, the changes in AT-BCL2 and AT-BAX were significantly correlated with each other, negatively correlated with the change in AUC insulin, and tended to be positively correlated with the change in circulating TNFα (Supplementary Figure SF1). We reason that the decrease in AT expression of these stress/death-related transducers, together with the downregulation of CCND1 and E2F3 (known effectors of cytokine signaling and cellular proliferation(32)), may represent an improvement in the basal metabolic and anti-inflammatory milieu in pioglitazone-treated subjects.

Although our study provides insight into novel mechanisms potentially driving and/or regulating the anti-diabetic effects of pioglitazone, several limitations need to be acknowledged. First, the relatively small sample size of the study is one key limitation that likely affected our power to detect significant changes in several of the measured features and clinical variables (hence our inability to conclusively reach significance for several relevant trends –i.e., in the changes of exosomal sorting-related genes and pro-apoptotic factors in AT). Second, although access to paired adipose tissue from the same study participants was very important to develop our working model, assessment of the differentially expressed miRNAs and their target genes in other tissues such as liver, pancreas, and muscle is required for a comprehensive assessment of EV-mediated mechanisms driving interorgan communication in response to anti-diabetic drugs. Finally, validation of our findings in an independent study cohort is necessary to confirm our results.

### Conclusions

In this study, we found that the levels of five miRNAs, namely miR-7-5p, miR-20a-5p, miR-92a-3p, miR-195-5p, and miR-374b-5p, were significantly downregulated in plasma EVs in response to pioglitazone treatment in people with T2D. Interestingly, downregulation of miR-195-5p in the plasma EVs contrasted with increased levels of the miRNA in AT. Downregulation of DICER and exosomal miRNA-sorting-related genes YBX1 and hnRNPA2B1 in AT suggests that miR-195-5p may accumulate in adipose tissue not because increased miRNA biogenesis but due to reduced exosomal sorting and disposal. Importantly, changes in the abundance of EV-miRNAs and AT-miRNAs correlated with changes in measures of glycemic control, β cell function, and lipid metabolism. These miRNAs overtargeted a network of transcripts that changed in AT in an apparently coordinated manner. Among key miRNA overtargeted and differentially expressed AT-genes, downregulated RAF1 may contribute to insulin-independent suppression of AT lipolysis, while downregulated CCND1 may contribute to enhanced mitochondrial function and lipogenesis in response to pioglitazone treatment. These events may contribute to the known pioglitazone-induced weight gain with concomitant improvement of insulin sensitivity and glucose homeostasis. Although additional studies are needed for confirmation, our study shows that some of the beneficial pharmacologic effects of pioglitazone may be effected by adipose-specific miRNA regulation and endocytic/exosomal trafficking.

## Methods

### Study Approval

All aspects of the study were approved by the Institutional Review Board of the University of Vermont. All study participants provided written informed consent prior to participation.

### Clinical trial design and subjects

Interventional study, ClinicalTrials.gov ID: NCT00656864 (33). Twenty-four participants with well-controlled type 2 diabetes (T2D) (HbA1C<7%) on diet and exercise or a stable dose of metformin were recruited for a randomized, double-blind, and placebo-controlled study. Subjects underwent a standard 75 g oral glucose tolerance test (OGTT) at baseline and after 12 weeks of treatment with either placebo (PLA, n=12) or pioglitazone (PIO, 45 mg/day; n=12). With the final sample size of 12 subjects in the active treatment group and 12 subjects in the control group, the study had over 80 % power to detect 1.5-fold change difference in miRNA expression using a two-sided test and significance level of 0.05. All aspects of the studies were approved by the Institutional Review Boards of the University of Vermont. All study participants provided written informed consent prior to participation.

### Blood analyses

Fasting blood samples were collected after an overnight fast in tubes containing EDTA or in serum-separating tubes. Plasma and serum samples were frozen at -80^?^C until analyses. HbA1C was measured on a COBAS INTEGRA 800 (Roche Diagnostics, Mannheim, Germany) automated analyzer. Insulin, C-peptide, and TNFα were measured by immunoassay (Insulin ELISA from Alpco, Salem, NH and Meso Scale Discovery, Rockville, MD, USA). Blood glucose concentrations were measured using YSI 2300 STAT Plus Glucose and L-Lactate Analyzer (YSI Incorporated, Yellow Springs, OH, USA). Data from the IVGTT were used to calculate insulin sensitivity (Si), acute insulin response to glucose (AIRg), and glucose effectiveness (Sg) using the Minimal Model method of Bergman (MINMOD-Millennium,© R. Bergman (34)). The IVGTT consisted of an intravenous bolus of 0.3 g dextrose/kg body weight followed by an insulin bolus (0.06 IU/kg body weight) 20 min later (33). Administration of the insulin bolus during the IVGTT circumvent the need for endogenous insulin secretion and permits the application of minimal model analysis in people with diabetes (35). Free fatty acids (FFA) were measured using the HR Series NEFA-HR(2) (Wako, Mountain View, CA) in vitro enzymatic colorimetric method for determination of non-esterified fatty acids in samples from the IVGTT, following the manufacturer’s instructions. The rate of FFA disappearance (K_FFA_) was calculated as the slope of the regression line fitting the natural logarithm of FFA concentration between 19 and 40 min, multiplied by -100 (36). This period was selected because it better represented, in our insulin-modified IVGTT experiment, the linear range of lipolysis suppression described for the second phase of FFA dynamics as described by Sumner et al. (37). The rate of FFA disappearance per unit of secreted insulin was calculated as reported by McLachlan et al. (36) “ as an insulin-corrected parameter, K_FFA/INS_, based on observations that insulin suppression of lipolysis is linear over the insulin dose range of 0–40 mU/l, and that interstitial concentration of insulin is within this range in the first 40 min of a 0.3-g/kg glucose IVGTT” (K_FFA/INS_ = K_FFA_ / incremental AUC_INS.0-40min_). Adipo-IR for each time point was calculated by multiplying the fasting FFA concentration (mmol/L) by the fasting insulin concentration (pmol/L) (38, 39).

### Adipose Tissue Biopsy

Subcutaneous periumbilical abdominal adipose tissue was obtained by percutaneous needle biopsy under local anesthesia. Tissue was aspirated into sterile PBS and washed twice to remove any blood. Whole tissue samples were then frozen in liquid nitrogen and stored at 280°C.

### Extracellular vesicles isolation and characterization

Extracellular vesicles were isolated from a total of 500 µl of fasting plasma samples using the Total Exosome Isolation Kit (from plasma) (ThermoFisher) according to the manufacturer’s instructions. Exosomal protein lysates were prepared with RIPA buffer with complete inhibitor cocktail (Roche Diagnostic) for protein quantification and Western blotting. Approximately 35 µg of exosomal protein was resolved in 4– 20% Criterion™ TGX™ Precast Midi Protein gels (Bio-Rad, Hercules, CA, USA), transferred to nitrocellulose membranes (Bio-Rad), and incubated overnight at 4°C with primary antibodies against CD9, CD63, and Calnexin. Membranes were then incubated in appropriate species-specific secondary antibodies for 1 h (IRDye 800CW anti-Rabbit IgG No. 926-32211 and IRDye 680RD anti-Mouse IgG No. 926-68070; Li-Cor Biosciences, Lincoln, NE). Protein bands were visualized using a Li-Cor Odyssey infrared imaging system (Li-Cor Biosciences). Quantification of extracellular vesicle concentration was conducted using the ExoELISA Complete Kit (CD9 Detection, calibrated using nanoparticle tracking analysis) from Systems Biosciences Inc (Palo Alto, CA) according to manufacturer’s instruction. Extracellular vesicle samples for Transmission Electron Microscopy (TEM) were fixed in 2% paraformaldehyde, deposited on Formvar-carbon coated electron microscopy grids and contrasted and embedded first with a solution of 2% uranyl acetate then in 0.4% uranyl acetate in 2% methyl cellulose (a mixture of 4% uranyl acetate and 2% methyl cellulose at a 1:9 ratio). TEM was performed using a FEI Morgagni TEM equipped with Gatan camera and microscope suite software. Nanoparticle Tracking Analysis (NTA) was performed with a NanoSight NS300 instrument and the NTA-3.4 software (Malvern Panalytical, Malvern). The instrument was equipped with a 488 nm blue laser module, flow-cell top plate, integrated temperature control, and a single-syringe pump module. Samples were diluted using cell culture grade water (Corning cat# 25-005-CI) to produce an optimal particle concentration for final measurement in the range of 10^7^ to 10^9^ particles/ml. Dilutions were initially assessed with a single quick static measurement of 30 second to identify the optimal dilution (which represented approximately 20 to 100 particles in the instrument’s field of view, per video frame). For final, more accurate quantification, 5 standard measurements of 1 minute of duration each were taken at a controlled temperature of 25 °C and under constant automatic flow (continuous syringe pump speed set to 50 arbitrary units). Camera level for video capture was set to 12 and detection threshold to 5 for all sample measurements.

### Profiling of miRNA abundance in circulating Evs

Total RNA from isolated extracellular vesicles was purified using Qiagen miRNeasy micro kit (Qiagen; Valencia, CA). miRNA expression was profiled by RT-PCR using custom designed 48-feature TaqMan MicroRNA Array cards (Life Technologies). Forty-two miRNAs, previously reported to be expressed in adipocyte-derived extracellular vesicles, were included in the custom panel. Raw Ct data was first adjusted based on the recovery of spike-in cel-miR-39, then normalized against the geometric mean of the top, most stable pair of miRNAs (miR-126 and miR-30b for this particular dataset) identified by the NormFinder algorithm. Features with less than 1.5-fold change were filtered out. Filtered log fold ratio data was used for differential expression analysis as described in the Statistical Analysis section.

### Expression profiling of select miRNAs and miRNA-overtargeted genes in human adipose tissue

Adipose tissue RNA was extracted using RNeasy Lipid Tissue Mini Kit (Qiagen) with RNase-Free DNase treatment (Qiagen). A cDNA library was made using Advantage-RT-for-PCR kits (Clontech; Mountain View, CA) using 1 μg RNA template and oligo-dT primers. Gene expression was measured by quantitative real-time PCR using TaqMan® reagents (TaqMan® assay list in Supplementary Table ST4) and ViiA-7® instrument from ThermoFisher Scientific (Waltham, MA), following the manufacturer’s instructions. Relative expression levels were calculated using the -ΔΔCt method. Expression data for adipose tissue miRNAs was normalized against endogenous control miR-191. Adipose tissue gene expression data was normalized against housekeeping gene PPIB. The -ΔΔCt data, equivalent to log fold ratio data, was used for differential expression analysis as described in the Statistical Analysis section.

### MicroRNA overtargeting network analysis

Our previous work studying disparate molecular mechanisms regulated by miRNAs indicated that system-wide miRNA-driven regulatory events commonly occur in a coordinated/cooperative fashion (14, 40-47). This process can be readily described by a compact network of interactions between relevant miRNAs and corresponding “ miRNA-overtargeted” genes (41, 43-46). The “ overtargeted” genes in a particular network appear to interact with a significantly higher number of differentially expressed miRNAs than expected by chance. In this study, we similarly conducted miRNA-overtargeting analyses following our published methodology (41, 43, 45, 46). Significance of the cooperative (more than 1 miRNA targeting a given gene) and overtargeting effect (significant higher number of miRNA-targeting events than expected by chance on a given gene) was assessed by comparing study-specific network proportions and the corresponding number of events against a collection of 10,000 simulated equivalent random networks. The list of validated targets supported by strong experimental evidence (i.e., reporter assay or Western blot) used for this analysis was downloaded from miRTarBase (file: miRTarBase_SE_WR.xls) using the SpidermiR package (48). Interaction networks were constructed using Cytoscape 3.5.1 (49). Enrichment of gene ontology annotations among sets of overtargeted genes was assessed using the GOCluster_Report function of the *systemPipeR* package (50) in the R environment.

### Statistical analysis

Differences in baseline clinical characteristics were assessed using the TableOne R package and the nonparametric Mann-Whitney U test (for continuous variables) or the Fisher exact test (for categorical variables). For assessment of longitudinal differences in clinical, metabolic, circulating miRNA, and AT-gene variables, mixed-effect models for repeated measures were implemented using the *nlme* R package. Measurements done in circulation were adjusted for body weight and hematocrit measurement to account for the potential confounding effects of weight gain and increased blood volume known to be induced by pioglitazone. Measurements of adipose tissue gene expression were only adjusted for body weight to account for the potential confounding effect of weight gain (the pioglitazone-induced increase in blood volume was considered to be a confounding factor only for variables measured in plasma or serum). Subject ID was included as a random effect variable to account for the correlations between repeated measurements within each patient. Partial correlations were also calculated in the R environment adjusting for the same covariates. Post-hoc analysis was performed using the *phia* package. Calculated effects and correlations with two-tailed P values < 0.05 were considered significant. False discovery rates (FDR) correcting for multiple testing were calculated using the Benjamini-Hochberg (BH) correction as implemented for the function *p.adjust()* in the *stats* R package. Considering that different investigators have different tolerance for false positives depending on the specific research purposes, no threshold for FDR was used. The FDR (BH-adjusted P value) is provided to give the readers an indication of the associated false discovery rate for each P value. Based on the well-established role of miRNAs in repression of target genes (51, 52), our statistical analysis of expression changes in miRNA-overtargeted genes in human adipose tissue *a priori* considered that upregulated AT-miRNAs would cause downregulation of overtargeted genes. Therefore, a one-sided P value < 0.05, was considered significant in those cases.

## Data Availability

All data produced in the present study are available upon reasonable request to the authors.

## Acknowledgments

The parent clinical trial was funded by Takeda Pharmaceuticals North America.

