## Supplementary Information for "Coordinated regulation of gene expression and microRNA changes in adipose tissue and circulating extracellular vesicles in response to pioglitazone treatment in humans with type 2 diabetes"

**Supplementary Table ST1**. Exosomal miRNA panel. Based on data reported by Thomou et al. 2017 and other references.

| **Idx** | **miRNA_ID** | **Thomou_CGL** | **Thomou_Mm** | **Tissue** | **References** | **TaqMan Cat #. 4427975-** | **Species** |
| --- | --- | --- | --- | --- | --- | --- | --- |
| 1 | cel-miR-39 | NA | NA | Spike_in_Control | Qiagen Instructions | 000200 | C. elegans |
| 2 | hsa-let-7f-5p | ns | significant | AT, Liv | Nunez Lopez et al. 2016, 2017; Wang et al. Gastroenterol 2012; Thomou et al. Nature 2017 | 000382 | conserved |
| 3 | hsa-miR-106b-5p | significant | significant | AT | Nunez Lopez et al. 2016, 2017; Thomou et al. Nature 2017 | 000442 | conserved |
| 4 | hsa-miR-107 | ns | significant | AT, Liv, Potential Control | Nunez Lopez et al. 2016, 2017; Vienberg et al. Acta Physiol 2017; Thomou et al. Nature 2017; Exiqon Guidelines 2015 | 000443 | conserved |
| 5 | hsa-miR-122-5p | ns | significant | AT | Nunez Lopez et al. 2016, 2017; Thomou et al. Nature 2017 | 002245 | conserved |
| 6 | hsa-miR-125b-5p | ns | ns | AT | Madam et al. Unpublished; Giroud et al. Mol Metabol 2016 | 000449 | conserved |
| 7 | hsa-miR-126-3p | significant | significant | AT | Nunez Lopez et al. 2016, 2017; Santovito et al JCEM 2014; Hubal et al. Obesity 2017; Thomou et al. Nature 2017 | 002228 | conserved |
| 8 | hsa-miR-133a-3p | significant | significant | AT | Vienberg et al. Acta Physiol 2017; Thomou et al. Nature 2017 | 002246 | conserved |
| 9 | hsa-miR-135b-5p | significant | significant | AT | Nunez Lopez et al. 2016, 2017; Thomou et al. Nature 2017 | 002261 | conserved |
| 10 | hsa-miR-138-5p | ns | significant | AT | Nunez Lopez et al. 2016, 2017; Thomou et al. Nature 2017 | 002284 | conserved |
| 11 | hsa-miR-149-5p | significant | significant | AT | Nunez Lopez et al. 2016, 2017; Thomou et al. Nature 2017 | 002255 | conserved |
| 12 | hsa-miR-152-3p | ns | significant | AT, Potential Control | Nunez Lopez et al. 2016, 2017; Thomou et al. Nature 2017 | 000475 | conserved |
| 13 | hsa-miR-155-5p | ns | significant | BAT | Madam et al. Unpublished; Vienberg et al. Acta Physiol 2017; Thomou et al. Nature 2017 | 002623 | conserved |
| 14 | hsa-miR-15a-5p | trend | significant | AT | Nunez Lopez et al. 2016, 2017; Thomou et al. Nature 2017 | 000389 | conserved |
| 15 | hsa-miR-16-5p | significant | significant | AT, Potential Control | Thomou et al. Nature 2017; Roberts et al PLoS One 2014 | 000391 | conserved |
| 16 | hsa-miR-181a-5p | ns | ns | AT, Liv, Endogenous Control | Wang et al. Gastroenterol 2012; Thomou et al. Nature 2017; Li et al Dis Markers 2015 | 000480 | conserved |
| 17 | hsa-miR-192-5p | significant | significant | AT | Nunez Lopez et al. 2016, 2017 | 000491 | conserved |
| 18 | hsa-miR-193b-3p | ns | significant | BAT | Madam et al. Unpublished; Vienberg et al. Acta Physiol 2017; Thomou et al. Nature 2017 | 002367 | mouse not listed? |
| 19 | hsa-miR-195-5p | significant | significant | AT | Santovito et al JCEM 2014; Kirby et al Physiol Genomics 2016; Thomou et al. Nature 2017 | 000494 | conserved |
| 20 | hsa-miR-196a-5p | significant | significant | BAT | Vienberg et al. Acta Physiol 2017; Thomou et al. Nature 2017 | 241070_mat | conserved |
| 21 | hsa-miR-199b-5p | significant | significant | AT, Liv | Wang et al. Gastroenterol 2012; Thomou et al. Nature 2017 | 000500 | mouse not listed? |
| **Idx** | **miRNA_ID** | **Thomou_CGL** | **Thomou_Mm** | **Tissue** | **References** | **TaqMan_Cat #. 4427975-** | **Species** |
| 22 | hsa-miR-203a-3p | ns | significant | BAT | Madam et al. Unpublished; Thomou et al. Nature 2017 | 000507 | conserved |
| 23 | hsa-miR-206 | significant | significant | AT | Nunez Lopez et al. 2016, 2017; Thomou et al. Nature 2017 | 000510 | conserved |
| 24 | hsa-miR-20a-5p | significant | significant | BAT | Madam et al. Unpublished; Thomou et al. Nature 2017 | 000580 | conserved |
| 25 | hsa-miR-21-5p | significant | significant | AT | Rodrigues et al. Cell Death Diff 2017; Thomou et al. Nature 2017 | 000397 | conserved |
| 26 | hsa-miR-221-3p | significant | significant | AT, Potential Control | Nunez Lopez et al. 2016, 2017; Thomou et al. Nature 2017; Li et al Dis Markers 2015 | 000524 | conserved |
| 27 | hsa-miR-222-3p | significant | significant | AT, Potential Control | Thomou et al. Nature 2017; Exiqon Guidelines 2015 | 002276 | conserved |
| 28 | hsa-miR-23a-3p | ns | significant | hemolysis, Endogenous Control | Thomou et al. Nature 2017; Exiqon Guidelines 2015 | 000399 | conserved |
| 29 | hsa-miR-26b-5p | ns | significant | BAT | Madam et al. Unpublished; Thomou et al. Nature 2017; Guller et al. BMC Genom 2015 | 000407 | conserved |
| 30 | hsa-miR-29c-3p | significant | significant | AT, Liv | Nunez Lopez et al. 2016, 2017; Thomou et al. Nature 2017 | 000587 | conserved |
| 31 | hsa-miR-30b-5p | significant | significant | BAT | Madam et al. Unpublished; Thomou et al. Nature 2017; Guller et al. BMC Genom 2015; Kim et al. JBC 2016 | 000602 | conserved |
| 32 | hsa-miR-320b | ns | significant | AT | Nunez Lopez et al. 2016, 2017; Thomou et al. Nature 2017 | 002844 | mouse not listed? |
| 33 | hsa-miR-325 | ns | significant | BAT | Thomou et al. Nature 2017 | 000540 | mouse not listed? |
| 34 | hsa-miR-328-3p | ns | significant | BAT | Madam et al. Unpublished; Oliverio et al. Nat Cell Bio 2016; Thomou et al. Nature 2017 | 000543 | conserved |
| 35 | hsa-miR-34c-3p | significant | ns | BAT, Liv | Madam et al. Unpublished; Thomou et al. Nature 2017; Chen et al. Nat Comm 2016 | 241009_mat | mouse not listed? |
| 36 | hsa-miR-365a-3p | significant | significant | BAT | Vienberg et al. Acta Physiol 2017; Thomou et al. Nature 2017 | 001020 | conserved |
| 37 | hsa-miR-374b-5p | significant | significant | AT | Thomou et al. Nature 2017; Santovito et al JCEM 2014; Kirby et al Physiol Genomics 2016 | 001319 | conserved |
| 38 | hsa-miR-375 | significant | significant | AT | Vienberg et al. Acta Physiol 2017; Thomou et al. Nature 2017 | 000564 | conserved |
| 39 | hsa-miR-378a-5p | significant | significant | BAT | Thomou et al. Nature 2017 | 000567 | conserved |
| 40 | hsa-miR-451a | ns | significant | hemolysis, Endogenous Control | Thomou et al. Nature 2017; Exiqon Guidelines 2015 | 001141 | conserved |
| 41 | hsa-miR-455-5p | significant | significant | BAT | Vienberg et al. Acta Physiol 2017; Thomou et al. Nature 2017 | 001280 | conserved |
| 42 | hsa-miR-7-5p | ns | trend | AT | Nunez Lopez et al. 2016, 2017; Thomou et al. Nature 2017 | 000268 | conserved |
| 43 | hsa-miR-92a-3p | ns | significant | BAT | Madam et al. Unpublished; Thomou et al. Nature 2017; Chen et al. Nat Comm 2016 | 000431 | mouse not listed? |
| 44 | hsa-miR-98-5p | ns | significant | AT | Thomou et al. Nature 2017 | 000577 | conserved |
| 45 | hsa-miR-99b-5p | ns | significant | AT | Thomou et al. Nature 2017 | 000436 | conserved |
| 46 | RNU44 | NA | NA | Endogenous Control | Madam et al. Unpublished | 001094 | mouse not listed? |
| 47 | RNU48 | NA | NA | Endogenous Control | Madam et al. Unpublished | 001006 | mouse not listed? |
| 48 | U6_snRNA | NA | NA | Endogenous Control | Madam et al. Unpublished | 001973 | conserved |

**Supplementary Table ST2**. Differential expression analysis of microRNAs in circulating exosomes and extracellular vesicles (collectively denominated EVs) **(A)** and in adipose tissue (AT) **(B)** from subjects with type 2 diabetes treated with pioglitazone, as compared to placebo controls. Data presented as log fold change (logFC) relative to placebo group baseline.

| (A) **EVs** | **PLACEBO (n=12)** | | **PIOGLITAZONE (n=12)** | | **Effect P Values** | | | **Effect FDR** | | |
| --- | --- | --- | --- | --- | --- | --- | --- | --- | --- | --- |
|  | **Baseline** | **Follow-up** | **Baseline** | **Follow-up** | **Group** | **Time** | **GxT** | **Group** | **Time** | **GxT** |
| **EV-miR-374-5p** | 0.0 (-4.6, 0.9) | 0.7 (-3.3, 2.1) | 0.7 (-1.7, 2.5) | -0.1 (-1.8, 2.3) | 0.1031 | 0.0032 | **0.0028** | 0.258 | 0.016 | **0.014** |
| **EV-miR-20a-5p** | 0.0 (-1.2, 1.5) | 0.6 (0.0, 1.2) | 0.4 (-0.4, 2.4) | 0.0 (-1.9, 0.7) | 0.4602 | 0.1471 | **0.0138** | 0.581 | 0.184 | **0.034** |
| **EV-miR-7-5p** | 0.0 (-4.4, 5.2) | 1.1 (-2.3, 6.5) | 2.4 (-1.5, 8.3) | 1.3 (-3.3, 3.1) | 0.0480 | 0.1283 | **0.0228** | 0.240 | 0.184 | **0.038** |
| **EV-miR-195-5p** | 0.0 (-1.4, 0.9) | 0.6 (-0.4, 1.1) | 0.2 (-1.6, 2.0) | -0.3 (-1.2, 0.9) | 0.8667 | 0.1359 | **0.0349** | 0.867 | 0.184 | **0.038** |
| **EV-miR-92a-3p** | 0.0 (-0.5, 1.6) | 0.4 (-0.3, 1.4) | 0.1 (-1.1, 2.8) | -0.1 (-3.9, 0.9) | 0.4648 | 0.9643 | **0.0377** | 0.581 | 0.964 | **0.038** |

| (B) **Adipose Tissue** | **PLACEBO (n=12)** | | **PIOGLITAZONE (n=12)** | | **Effect P Values** | | | **Effect FDR** | | |
| --- | --- | --- | --- | --- | --- | --- | --- | --- | --- | --- |
|  | **Baseline** | **Follow-up** | **Baseline** | **Follow-up** | **Group** | **Time** | **GxT** | **Group** | **Time** | **GxT** |
| **AT-miR-195-5p** | 0.0 (-0.5, 0.7) | -0.3 (-0.6, 0.1) | -0.2 (-0.8, 0.3) | 0.1 (-0.7, 0.6) | 0.1624 | 0.0297 | **0.0063** | 0.735 | 0.043 | **0.032** |
| **AT-miR-7-5p** | 0.0 (-1.7, 2.0) | 0.0 (-1.6, 0.7) | -0.3 (-2.2, 0.4) | 0.0 (-1.0, 3.7) | 0.4409 | 0.6831 | 0.1133 | 0.735 | 0.683 | 0.283 |
| **AT-miR-374b-5p** | 0.0 (-1.0, 0.9) | -0.7 (-2.5, 0.4) | -0.6 (-1.4, 0.6) | -0.5 (-2.4, 0.4) | 0.3120 | 0.0317 | 0.1983 | 0.735 | 0.043 | 0.311 |
| **AT-miR-20a-5p** | 0.0 (-1.1, 1.0) | -0.8 (-1.1, 0.6) | -0.2 (-1.1, 0.8) | -0.4 (-1.2, 0.6) | 0.8110 | 0.0268 | 0.2485 | 0.984 | 0.043 | 0.311 |
| **AT-miR-92a-3p** | 0.0 (-0.7, 2.1) | 0.8 (0.0, 2.2) | 0.2 (-0.9, 1.0) | 0.8 (-0.2, 2.1) | 0.9843 | 0.0347 | 0.7971 | 0.984 | 0.043 | 0.797 |

**Supplementary Table ST3**. Pair-wise partial correlations (adjusted for body weight) among -differentially expressed miRNA-overtargeted genes in adipose tissue (AT).

| **feature1.change** | **feature2.change** | **n** | **r** | **p.value** | **FDR** |
| --- | --- | --- | --- | --- | --- |
| DICER | BCL2 | 18 | 0.84 | 4.24E-06 | 0.00024032 |
| RAF1 | BCL2 | 18 | 0.81 | 1.23E-05 | 0.000596889 |
| DICER | BAX | 18 | 0.81 | 1.47E-05 | 0.000625861 |
| DICER | CCND1 | 18 | 0.81 | 1.78E-05 | 0.000672438 |
| YBX1 | CCND1 | 18 | 0.79 | 3.44E-05 | 0.00116799 |
| BCL2 | BAX | 18 | 0.76 | 0.000105069 | 0.002423834 |
| hnRNPA2B1 | CCND1 | 18 | 0.73 | 0.000267066 | 0.003947929 |
| BCL2 | CCND1 | 18 | 0.69 | 0.000708441 | 0.00766578 |
| CCND1 | BAX | 18 | 0.68 | 0.000996926 | 0.009160946 |
| BCL2 | E2F3 | 18 | 0.64 | 0.002620004 | 0.018947237 |
| RAF1 | BAX | 18 | 0.63 | 0.002704549 | 0.018947237 |
| BAX | E2F3 | 18 | 0.62 | 0.00344438 | 0.021292531 |
| DICER | E2F3 | 18 | 0.62 | 0.003745654 | 0.022657245 |
| CCND1 | E2F3 | 18 | 0.56 | 0.010846697 | 0.044432253 |
| DICER | RAF1 | 18 | 0.55 | 0.012497879 | 0.046842694 |
| YBX1 | BCL2 | 18 | 0.54 | 0.013618645 | 0.048740413 |

**Supplementary Table ST4.** Individual TaqMan® Assays used for quantification of adipose tissue miRNAs and miRNA-overtargeted genes.

TaqMan® Assay Name

TaqMan Assay ID: 000268 (hsa-miR-7-5p)

TaqMan Assay ID: 000580 (hsa-miR-20a)

TaqMan Assay ID: 000431 (hsa-miR-92a-3p)

TaqMan Assay ID: 002299 (hsa-miR-191-5p)

TaqMan Assay ID: 000494 (hsa-miR-195-5p)

TaqMan Assay ID: 001319 (hsa-miR-374b-5p)

TaqMan Assay ID: Hs01084653_m1 (AGO1)

TaqMan Assay ID: Hs00293044_m1 (AGO2)

TaqMan Assay ID: Hs04259596_g1 (AKR7A2)

TaqMan Assay ID: Hs03044164_m1 (BAMBI)

TaqMan Assay ID: Hs99999001_m1 (BAX)

TaqMan Assay ID: Hs00608023_m1 (BCL2)

TaqMan Assay ID: Hs00197982_m1 (BCL211)

TaqMan Assay ID: Hs00176148_m1 (BMPR2)

TaqMan Assay ID: Hs00234387_m1 (CASP3)

TaqMan Assay ID: Hs00765553_m1 (CCND1)

TaqMan Assay ID: Hs01026536_m1 (CCNE1)

TaqMan Assay ID: Hs00229023_m1 (DICER1)

TaqMan Assay ID: Hs00605457_m1 (E2F3)

TaqMan Assay ID: Hs00924627_g1 (FLNA)

TaqMan Assay ID: Hs00242600_m1 (HNRNPA2B1)

TaqMan Assay ID: Hs00275843_s1 (IRS2)

TaqMan Assay ID: Hs00206553_m1 (KIAA0100)

TaqMan Assay ID: Hs00559840_m1 (KRT7)

TaqMan Assay ID: Hs01558224_m1 (MAPK9)

TaqMan Assay ID: Hs00168719_m1 (PPIB)

TaqMan Assay ID: Hs00234119_m1 (RAF1)

TaqMan Assay ID: Hs00186212_m1 (RGS5)

TaqMan Assay ID: Hs00998193_m1 (SMAD7)

TaqMan Assay ID: Hs00920354_m1 (SMPD3)

TaqMan Assay ID: Hs00299877_m1 (TMEM97)

TaqMan Assay ID: Hs00900055_m1 (VEGFA)

TaqMan Assay ID: Hs02742755_g1 (YBX1)

TaqMan Assay ID: Hs01119384_g1 (WEE1)

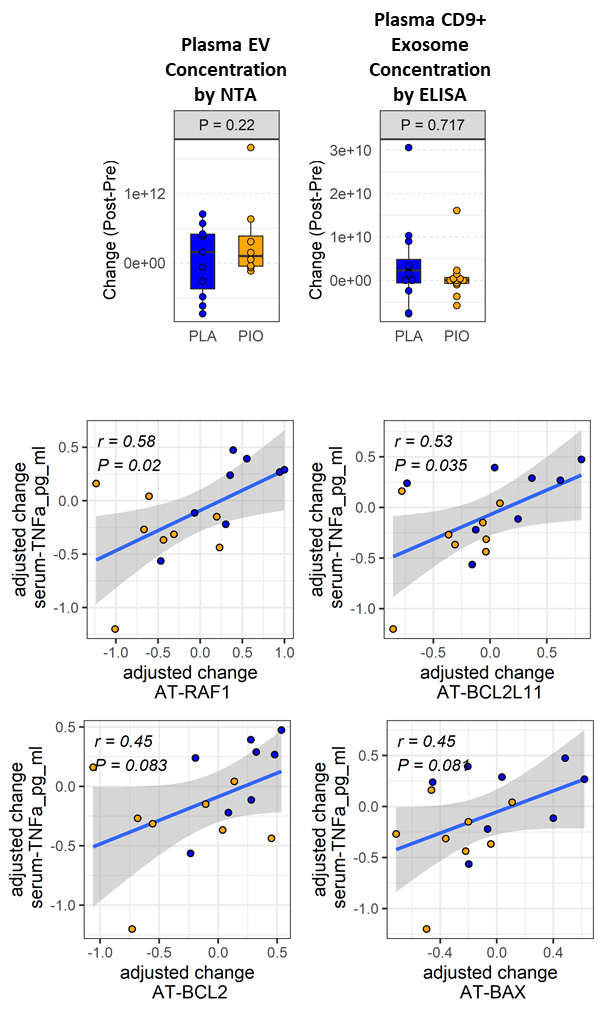

**Supplementary Figure SF1**. Quantification of EVs by nanoparticle tracking analysis (NTA) and CD9+ exosomes by ELISA (A,B). Partial correlations between the change in AT genes and the change in serum TNFα.
